# Breaking the Growth Barrier: Stunting Among Afghan Children (6–59 Months) – Insights from the 2023 UNICEF MICS Survey

**DOI:** 10.1101/2025.01.21.25320890

**Authors:** Mokhtar Ashor, MD. Sajid Sultan Haque, Shagufta Baig

**Affiliations:** James P Grant School of Public Health, BRAC University, Dhaka, Bangladesh

## Abstract

Stunting has emerged as a significant health problem in South Asia. Afghanistan was hit the hardest with the highest prevalence of stunting in 2018 in the region. With the recent education ban for women and girls, Afghanistan has become prone to even worse satiation. Additionally, with respect to the country’s social and economic profile after the COVID-19 pandemic, the country and its population, specifically women and children, faced challenges in receiving essential living services. The objective of this study was to determine the current prevalence of stunting and the factors that are significantly associated with this disease among children.

The study uses the MICS nationally representative dataset in which the survey was conducted from 2022--2023 in Afghanistan by UNICEF. The results revealed a high prevalence rate of stunting among children aged 6--59 months (48%), and several factors (mother and caregiver education, household wealth, children’s age and infectious diseases) were associated with stunted children in the country. The major part of the country in western Afghanistan has more children suffering from stunting than the other parts of the country do.

Our findings highlight the importance of mothers’ education in preventing their children from stunting, and our research recommends that the government focus on interventions in the community to increase awareness and make nutritional services available to people in hard-to-reach areas of the country.

## Introduction

Malnutrition remains one of the most pressing global public health challenges, disproportionately affecting vulnerable populations such as children, women, and those in low-income and conflict prone countries. Globally, undernutrition contributes to nearly half of all deaths in children under five years of age, emphasizing the severity of this issue. Malnutrition can be defined as undernutrition and overnutrition, in which cases can involve excessive or low intake of macro- and micronutrients, which are necessary for the body to take in balance to function well (1).The dual burden of malnutrition—where undernutrition coexists with overnutrition—further complicates efforts to address nutritional disparities, especially in regions experiencing rapid economic and social transitions. In this study, we aimed to look for undernutrition only, which we can name these types of undernutrition: stunting (defined as low height over age), wasting (defined as low weight for height) and underweight (low weight for age) (1).

Stunting is a type of chronic malnutrition that remains a major problem for global health and affects millions of children worldwide; more particularly, the number of affected children is large in low- and middle-income countries (2). To what extent stunted children show growth reflects or physically underdevelopment, the consequences are not limited here; according to the WHO, it also impairs cognitive and motor development, which can affect the whole society in the long term. Globally, approximately 45% of deaths among U5 children are caused by undernutrition (3). The data revealed that more than 148.1 million children aged U5 were stunted in 2023, and a large portion of the data fall in areas where low- and middle-income countries are located (WHO) (1).

Around the world, sub-Saharan Africa and South Asia face the highest prevalence of stunting, which is not an evenly distributed burden of the disease (4, 5). South Asia alone shows that 33% of the children in these areas are stunted due to a lack of access to healthcare, inadequate sanitation, poor maternal health, food security and mostly poverty (6). Despite the improvements made in this area in recent years, the pace of reduction in children’s health, specifically stunting, has remained insufficient to reach global targets, stimulating further research into the specific characteristics of this condition in diverse regional contexts (7).

Afghanistan, a country characterized by decades of conflict, economic instability, and environmental challenges, has one of the highest rates of childhood stunting in the world. Factors such as food insecurity, poor maternal health, inadequate breastfeeding practices, and limited access to clean water and sanitation have contributed to this crisis (8). Considering all the other covariant factors, the prevalence of stunting is assumed to be much greater than previously reported, which shows that approximately 40% of children were stunted around the country in 2018 according to the Afghanistan Health Survey, which already makes Afghanistan one of the countries with the highest rate of stunting in the world (9). To a greater extent, the factors that have the greatest effect are widespread poverty, food insecurity, conflict, inadequate access to healthcare, nutritional services and natural disasters, which have recently significantly undermined efforts to improve food security and maternal and child health (10).

The complex nature of the disease and insufficient in-depth data on the causes of stunting, together with geographical barriers, make it difficult to address it in Afghanistan, even with multiple interventions at the national and international levels (11). Previous studies also highlighted the number of causes related to stunting, including household number, income, lack of nutritional diversity and limited access to sanitary facilities and clean water (12, 13). To better understand the risk factors for stunting in countries, however, comprehensive and context-specific research is desperately needed, especially given the nation’s distinct sociopolitical and environmental conditions. However, to close this gap, this study looks at UNICEF’s 2023 Multiple Indicator Cluster Survey (MICS) to check how common stunting is in Afghan children aged 6--59 months. Many factors, such as social, economic, and environmental factors, may lead to stunting, and all these factors can be examined with the MICS data.

## Methods

### Data Source

The study utilized data from the most recent UNICEF Multiple Indicator Cluster Survey (MICS) data, which were obtained from 2022–2023. In the survey for the selection of the sample, a multistage, stratified cluster sampling approach was used. For the initial phase, the sampling frame was based on the 2019 satellite imagery-based frame. The primary sampling units (PSUs) selected at the first stage were the enumeration areas (EAs) defined for the census enumeration. Each sample EA underwent a new listing of households, and in the second step, a sample of households was chosen. This is a type of probability sample where after the list of households in the sample PSUs and the list of household members in each interviewed sample household is full, each household and household member has a positive and known probability of being selected. By weighting the data by the inverse of the overall probabilities of selection, probability sampling allows for valid inference of the population or any subgroup of the population. A full explanation of the sampling design can be found in the UNICEF MICS survey report 2022--2023 (14).

For this study, the study population is the total number of children aged 6--59 months whose mothers or one of their caregivers were interviewed in this survey, and the anthropometric measurements were taken by the survey team to record their height, weight, and middle and upper arm circumstances (14).

### Outcome variables

The primary outcome variable was stunting among the children aged 6-59 Months. Children defined as stunted were those whose height-for-age z score was less than -2 according to the WHO standard. The children were weighed, and their height was measured carefully during the interviews by the survey team. For our sample, we excluded children who were younger than 6 months, and the final sample size for this study was 29.552. We then calculated the z score for all the children included in this study and categorized them as stunted children if they fell below the -2 score and not stunted children if they were above the - 2 z score.

### Covariates

We took into consideration a number of factors in our multivariate model analyses, primarily to control for potential confounders that might affect the study’s conclusions. The factors included the children’s age distribution, the mother’s or caregiver’s greatest level of education, the children’s place of residence, the size of the household (number of members), and the household wealth index (which was divided into quintiles, from the lowest to the richest). Using data on household assets supplied by the survey authority, principal component analysis (PCA) was used to create the wealth index. By using this approach, the wealth index is guaranteed to fairly represent the socioeconomic standing of the households included in the research. Our attention to these factors was intended to guarantee that the analysis was sound and that these important demographic factors did not skew the results.

### Statistical analysis

Given the complex design of the MICS survey, survey weights were applied prior to analysis to account for the sampling design and ensure that the results were representative of the population. The prevalence of stunting among children aged 6–59 months was calculated and reported according to various background characteristics while considering the survey’s complex design and weighted data. Additionally, age- and sex-standardized rates of stunting were computed to reduce the influence of demographic differences among participants. Descriptive analyses were followed by regression modeling to explore the associations between explanatory variables and stunting. Both univariate and multivariate logistic regression analyses were conducted to identify factors independently associated with stunting.

With a significance level set at p < 0.05 for adjusted analyses. The results are presented as prevalence ratios (PRs), 95% confidence intervals (CIs), and p values. Data analysis was performed via STATA version 17.

### Results

The results in Table 1 show that the research included 29,552 children aged 6 to 59 months. The largest age group is 37--59 months, accounting for 44.7% of the children. The smallest group ranged from 6--12 months, at 12.83%. When we look at gender, 51.02% of the children are boys, and 48.98% are girls. The children are spread across eight regions. The central area (Kabul, Parwan, Kapisa, Panjshir, Maidan Wardak, Logar) has the greatest percentage, at 16.88%, and the central highlands (Bamyan, Daykundi) have the lowest percentage, at 4.61%. A large percentage of the children, 84.98%, live in rural areas of the country, and only 15.02% live in cities. In terms of mothers’ education level, 83% of them had no school education or had just gone to preprimary education. Only 2.34% of the mothers had gone to higher school. In terms of the wealth index, 24.06% of the children are from the poorest homes, whereas 12.25% are from the richest homes. With respect to health conditions, 37.82% of the children had diarrhea, whereas 61.92% did not. A small percentage (0.01%) did not reply to the question concerning diarrhea.

**Table 1.** Sociodemographic characteristics of the study population (N = 29,552)

| **Characteristic** | **N (%)** |
| --- | --- |
| **Age (Month)** |  |
| 6-12 | 3,791 (12.83) |
| 13-24 | 6,361 (20.94) |
| 25-36 | 6,361 (21.52) |
| 37-59 | 13,211 (44.7) |
| **Child Sex** |  |
| Male | 15,076 (51.02) |
| Female | 14,476 (48.98) |
| **Regions** |  |
| Central | 4,989 (16.88) |
| North | 4,111 (13.91) |
| South East | 3,988 (13.49) |
| Central Highlands | 1,361 (4.61) |
| East | 3,988 (13.48) |
| West | 3,161 (10.7) |
| North East | 2,987 (10.11) |
| South | 4,970 (16.82) |
| **Residence** |  |
| Urban | 4,440 (15.02) |
| Rural | 25,112 (84.98) |
| **Mother's Education** |  |
| Pre-Primary/ECE or None | 24,527 (83.00) |
| Primary | 2,203 (7.45) |
| Lower Secondary | 929 (3.14) |
| Upper Secondary | 1,201 (4.06) |
| Higher | 692 (2.34) |
| **Wealth index** |  |
| Poorest | 7,110 (24.06) |
| Poorer | 7,017 (23.74) |
| Middle | 6,495 (21.98) |
| Richer | 5,310 (17.97) |
| Richest | 3,620 (12.25) |
| **Child Had Diarrhea** |  |
| Yes | 11,177 (37.82) |
| No | 18,298 (61.92) |
| Don't Know | 74 (0.25) |
| No Response | 3 (0.01) |

The prevalence of stunting among the children was 48% (Figure 1), with a significant association with the age of the children, area of residency, education level of the child’s mother and caregiver, wealth index and children suffering from diarrhea (all P < 0.001) (Table 2).

**Table 2.** Associations between sociodemographic characteristics and stunting.

| Characteristic | Overall | Stunted | Not Stunted | p value |
| --- | --- | --- | --- | --- |
| **Age** |  |  |  |  |
| 6-12 | 3,791 (12.83) | 986 (7.00) | 2,805 (18.14) | <0.001 |
| 13-24 | 6,189 (20.94) | 2,739 (19.44) | 3,450 (22.31) |  |
| 25-36 | 6,361 (21.52) | 3,508 (24.90) | 2,853 (18.45) |  |
| 37-59 | 13,211 (44.70) | 6,853 (48.65) | 6,358 (41.11) |  |
| **Regions** |  |  |  |  |
| Central | 4,989 (16.88) | 1,973 (14.01) | 3,016 (19.50) | <0.001 |
| North | 4,111 (13.91) | 1,843 (13.08) | 2,268 (14.66) |  |
| South East | 3,988 (13.49) | 1,791 (12.71) | 2,197 (14.21) |  |
| Central Highlands | 1,361 (4.61) | 552 (3.92) | 809 (5.23) |  |
| East | 3,985 (13.48) | 2,061 (14.63) | 1,924 (12.44) |  |
| West | 3,161 (10.70) | 1,640 (11.64) | 1,521 (9.83) |  |
| North East | 2,987(10.11) | 1,416 (10.05) | 1,571 (10.16) |  |
| South | 4,987 (16.82) | 2,810 (19.95) | 2,160 (13.97) |  |
| **Residence** |  |  |  |  |
| Urban | 4,440 (15.02) | 1,592 (11.30) | 2,848 (18.41) | <0.001 |
| Rural | 25,112 (84.98) | 12,494 (88.70) | 12,618 (81.59) |  |
| **Mother's Education** |  |  |  |  |
| Pre-Primary/ECE or None | 24,527 (83.00) | 12,290 (87.25) | 12,237 (79.12) | **<0.001** |
| Primary | 2,203 (7.45) | 871 (6.18) | 1,332 (8.61) |  |
| Lower Secondary | 929 (3.14) | 322 (2.29) | 607 (3.92) |  |
| Upper Secondary | 1,201 (4.04) | 418 (2.97) | 783 (5.06) |  |
| Higher | 692 (2.34) | 185 (1.31) | 507 (3.28) |  |
| **Wealth index** |  |  |  |  |
| Poorest | 7,110 (24.06) | 3,980 (28.26) | 3,130 (20.24) | **<0.001** |
| Poorer | 7,017 (23.74) | 3,697 (26.25) | 3,320 (21.47) |  |
| Middle | 6,495 (21.98) | 3,162 (22.45) | 3,333 (21.55) |  |
| Richer | 5,310 (17.97) | 2,167 (15.38) | 3,143 (20.32) |  |
| Richest | 3,620 (12.25) | 1,080 (7.67) | 2,540 (16.42) |  |
| **Child Had Diarrhea** |  |  |  |  |
| Yes | 11,177 (37.82) | 5,474 (38.86) | 5,701 (36.87) | **0.002** |
| No | 18,298 (61.92) | 8,581 (60.92) | 9,717 (62.83) |  |
| Don't Know | 74 (0.25) | 29 (0.21) | 45 (0.29) |  |
| No Response | 3 (0.01) | 2 (0.01) | 1 (0.01) |  |

**Figure 1.**
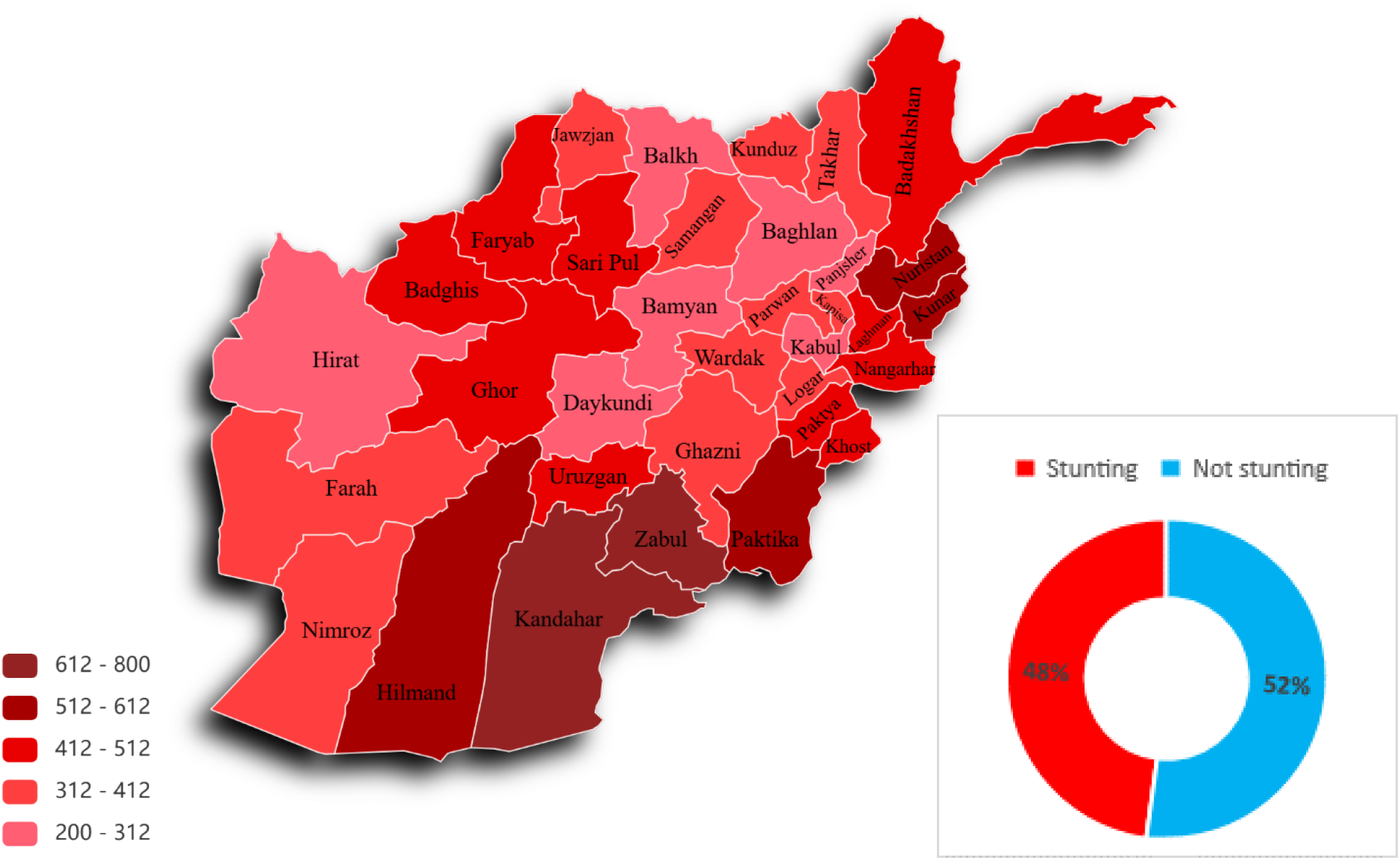
The prevalence of stunting among children aged 6--59 months in Afghanistan. The prevalence of stunting among children aged 6--59 months is 48%, as the country heat map shows that the number of stunted children is high in the western region of the country.

According to our univariate and multivariate analyses (Table 3), when children are aged 6--12 months, older children aged 13--24, 25--36, and 37--59 months are more likely to be too short for their age (stunting). As children grow older, their odds of being stunted increase with age, with children aged 25--36 months having the highest chance (AOR = 3.55; 95% CI: 3.25, 3.88; P <0.001). Additionally, children from rural parts of the country are more likely to be too short for their age than are those from urban areas (AOR = 1.17; 95% CI: 1.08, 1.26; P<0.001), even though this effect decreases when adjustments are made for other mixed factors. A high level of mom or child caretaker education is associated with a lower chance of being stunted. As education level increases from low to high, the chance of being too short decreases substantially, and mothers with higher education have an AOR of 0.56 (95% CI: 0.47, 0.67) compared with those with no or preprimary education. Wealthier households are associated with significantly lower odds of stunting. Compared with those from the poorest households, children from the richest quintile are much less likely to be stunted (AOR = 0.40; 95% CI: 0.37, 0.45 P<0.001).

**Table 3.**
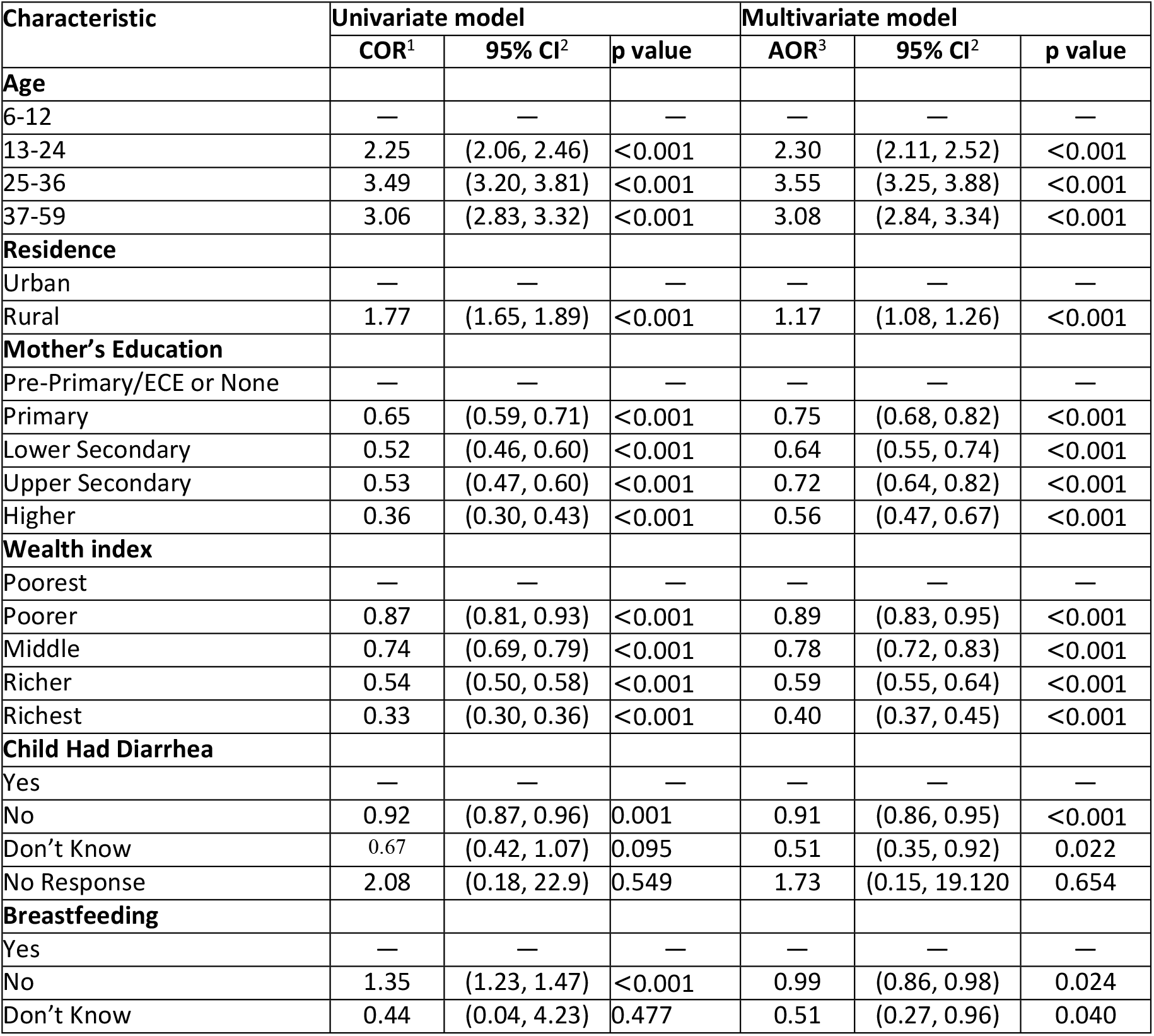
Univariate and multivariate logistic regression analyses of factors associated with stunting among children aged 6–59 months in Afghanistan.

| Characteristic | Univariate model |  |  | Multivariate model |  |  |
| --- | --- | --- | --- | --- | --- | --- |
|  | COR<sup>1</sup> | 95% CI<sup>2</sup> | p value | AOR<sup>3</sup> | 95% CI<sup>2</sup> | p value |
| **Age** |  |  |  |  |  |  |
| 6-12 | — | — | — | — | — | — |
| 13-24 | 2.25 | (2.06, 2.46) | <0.001 | 2.30 | (2.11, 2.52) | <0.001 |
| 25-36 | 3.49 | (3.20, 3.81) | <0.001 | 3.55 | (3.25, 3.88) | <0.001 |
| 37-59 | 3.06 | (2.83, 3.32) | <0.001 | 3.08 | (2.84, 3.34) | <0.001 |
| **Residence** |  |  |  |  |  |  |
| Urban | — | — | — | — | — | — |
| Rural | 1.77 | (1.65, 1.89) | <0.001 | 1.17 | (1.08, 1.26) | <0.001 |
| **Mother's Education** |  |  |  |  |  |  |
| Pre-Primary/ECE or None | — | — | — | — | — | — |
| Primary | 0.65 | (0.59, 0.71) | <0.001 | 0.75 | (0.68, 0.82) | <0.001 |
| Lower Secondary | 0.52 | (0.46, 0.60) | <0.001 | 0.64 | (0.55, 0.74) | <0.001 |
| Upper Secondary | 0.53 | (0.47, 0.60) | <0.001 | 0.72 | (0.64, 0.82) | <0.001 |
| Higher | 0.36 | (0.30, 0.43) | <0.001 | 0.56 | (0.47, 0.67) | <0.001 |
| **Wealth index** |  |  |  |  |  |  |
| Poorest | — | — | — | — | — | — |
| Poorer | 0.87 | (0.81, 0.93) | <0.001 | 0.89 | (0.83, 0.95) | <0.001 |
| Middle | 0.74 | (0.69, 0.79) | <0.001 | 0.78 | (0.72, 0.83) | <0.001 |
| Richer | 0.54 | (0.50, 0.58) | <0.001 | 0.59 | (0.55, 0.64) | <0.001 |
| Richest | 0.33 | (0.30, 0.36) | <0.001 | 0.40 | (0.37, 0.45) | <0.001 |
| **Child Had Diarrhea** |  |  |  |  |  |  |
| Yes | — | — | — | — | — | — |
| No | 0.92 | (0.87, 0.96) | 0.001 | 0.91 | (0.86, 0.95) | <0.001 |
| Don't Know | 0.67 | (0.42, 1.07) | 0.095 | 0.51 | (0.35, 0.92) | 0.022 |
| No Response | 2.08 | (0.18, 22.9) | 0.549 | 1.73 | (0.15, 19.120) | 0.654 |
| **Breastfeeding** |  |  |  |  |  |  |
| Yes | — | — | — | — | — | — |
| No | 1.35 | (1.23, 1.47) | <0.001 | 0.99 | (0.86, 0.98) | 0.024 |
| Don't Know | 0.44 | (0.04, 4.23) | 0.477 | 0.51 | (0.27, 0.96) | 0.040 |

Children who did not experience diarrhea had a slightly lower likelihood of stunting (AOR = 0.91; 95% CI: 0.86, 0.95; P<0.001) than did those who did experience diarrhea. The results from the univariate analysis revealed that children who were not breastfed had greater odds of being stunted than did children who were breastfed (AOR=1.35 CI: 1.23, 1.47; P<0.001).

## Discussion

This research revealed a high prevalence rate of stunting (48%) among children aged 6–59 months in Afghanistan, which is similar to the findings of previous studies from the same context in low- and middle-income countries (15, 16). Our analysis revealed significant associations, such as sociodemographic factors, including child age, residency area, maternal education, family wealth status and diarrhea with child stunting. These findings indicate that stunting is influenced by both socioeconomic and health-related factors.

Our research indicates that the risk of stunting increases as a child grows, with children aged 25–36 months having the greatest effect. This phenomenon has also been underscored in the previous literature, suggesting that stunting becomes increasingly intense as children grow older because of prolonged exposure to malnutrition, chronic diseases, and insufficient diversity (10, 17). Youth-centric interventions, especially throughout the 1st 1,000 days of life, might be important in halting stunting progression as children age (18). Studies have also demonstrated that there is a differentiation in severe hindering between children from rural areas and those from urban areas, which indicates that children in rural areas are at a greater stake, although the amount is reduced when other factors are taken into account (19, 20, 21, 22). Other evidence suggests that in rural areas, undernutrition rates, with an emphasis on chronic undernutrition, are higher than they are in urban areas, with rural children often suffering from more deprived nutritional conditions owing to restricted access to healthcare services, limited dietary diversity and poor environmental sanitation and cleanliness (23, 24). To make significant contributions to the worldwide epidemic of stunting, it is crucial to confront these structural constraints, focusing intentionally on essential interventions for rural areas.

Maternal education was found to be one of the strongest protective factors against stunting. Mothers with higher education levels had significantly lower odds of having stunted children, even when adjustments were made for other variables. A mother’s education could enable her to offer her children better nutrition and health care and equip her with appropriate knowledge for healthy feeding and disease prevention (25, 26). This underscores how important it is when trying to reduce child malnutrition to intervene in maternal education. The stunting is strongly correlated with the wealth index; the children from the richest households are at a much lower risk of being stunted than those in the poorest quintile are, as wealth is a proxy for the ability of a household to meet the nutritional needs of its members. Other studies also reported a positive correlation between household wealth and stunted children (27, 28).

The study revealed that diarrhea is a significant risk factor for stunting since children with no history of diarrhea are less likely to be stunted. In general, diarrheal diseases are responsible for the prevailing malnutrition in communities by impairing nutrient absorption and increasing nutrient losses (29). The high prevalence of diarrhea in children in the study areas suggests that improving water, sanitation and hygiene practices alongside health care has the potential to significantly reduce stunting rates. Although univariate analysis revealed that children who did not receive breastmilk were at increased risk of stunting, it was eliminated from the multivariable model. It can be interpreted from this observation that merely practicing just breastfeeding is not enough to guard off stunting, and it can also be that most children do not breastfeed after 2 years of age; alternatively, there are other factors, such as maternal education and household wealth, which have a greater influence on the condition. However, breastfeeding remains a key practice in overall child health promotion, and future studies may want to uncover its relationship with stunting in a more in-depth manner while accounting for other confounders.

## Strengths and Limitations of the Study

However, this study provides valuable insight into the factors associated with stunting, considering its limitations due to its design as a cross-sectional study, which limits the ability to create a cause-and-effect relationship. Moreover, the data used in this study are based on self-reported questions, which cannot prevent recall bias. However, the implications of our findings can be significant for public health. In turn, focused programs to eliminate stunting should include not only interventions to increase maternal education, which is a crucial factor in and of itself but also in such priority areas as improving access to healthcare in rural settings and addressing root problems of social inequity that fuel malnutrition.

## Conclusion

In summary, this research shows that 48% of Afghan children aged 6--59 months are too short for their age (stunting). The main reasons are the child’s age, residency area, maternal education level, household economic status, and diarrhea. Children in rural areas of the country, particularly those with mothers who have low levels of education, are often shorter than expected for their age. Older children, aged 25–36 months, were at the greatest risk. By addressing these underlying causes, Afghanistan can make strides in reducing the prevalence of stunting and improving the health and well-being of its youngest population. These efforts are critical for breaking the intergenerational cycle of malnutrition and poverty, aligning with global goals such as Sustainable Development Goals 2 and 3 to end hunger and promote good health. Therefore, as these research results emphasize, we need plans for better education of child mothers, a reduction in being poor, and better health care to fight stunting in Afghanistan.

## Data Availability

MICS UNICEF is the upholder of the 2022--2023 MICS data, which are freely available upon reasonable request submission to MICS-UNICEF.

https://www.unicef.org/afghanistan/reports/afghanistan-multiple-indicator-cluster-survey-mics-2022-2023

## Acknowledgments

We acknowledge the Multi Indicator Cluster Survey (MICS – UNICEF) for approving the MICS Afghanistan data.

## Authors’ contributions

**MA**: Conceptualized the study, designed the methodology, conducted data analysis and interpretation and drafted the initial manuscript. **SSH:** Assisted in the study design and contributed to data cleaning process. Provided critical input on the analysis and interpretation of results. Reviewed and edited the manuscript for intellectual content. **SB:** Contributed to literature review and prepared the initial draft of the introduction and discussion sections. Supported manuscript formatting and ensured adherence to journal guidelines. All the Author’s reviewed and approved the final manuscript.

## Ethics approval and norms

The survey protocol, which was approved by the NSIA Technical Committee in July 2022, adhered to internationally recognized ethical guidelines, including the UNICEF Ethical Guidelines and Afghanistan’s National Ethical Guidelines **–** The Ministry of Public Health (MoPH). The protocol also included a Protection Protocol that outlines potential risks during the survey lifecycle and management strategies to mitigate these risks, ensuring the ethical treatment of participants. Written consent was obtained for each respondent participating and adult consent was obtained in advance of the child’s assent. All respondents were informed of the voluntary nature of participation and the confidentiality and anonymity of information. Additionally, respondents were informed of their right to refuse to answer all or questions, as well as to stop the interview at any time.

Before the process of writing and analyzing the nationally representative secondary dataset of the MICS 2022-2023 survey dataset, the official written permission was obtained through email to use the dataset for further analysis.

## Consent for publication

Consent was obtained from the MICS-UNICEF.

